# Could Metformin use reduce abdominal aortic aneurysm risk? A Mendelian randomisation study using known Metformin targets

**DOI:** 10.1101/2023.10.13.23293381

**Authors:** Katie L Saxby, Frank Dudbridge, Nilesh J Samani, Matthew J Bown, Christopher P Nelson

## Abstract

**Introduction:** An abdominal aortic aneurysm (AAA) is a swelling of the main artery in the body estimated to affect 0.92% of adults (aged 30-79) worldwide. Rupture is often fatal and surgical intervention may be offered if the risk of rupture is high. There is no treatment to prevent AAA or to slow aneurysm growth aside from dietary and lifestyle recommendations. Metformin, a drug prescribed to treat type 2 diabetes, has previously been associated with a potential reduction in AAA disease risk but no causal link has been shown. Here we investigate the causal link between Metformin and AAA risk through Mendelian randomisation (MR).

**Methods:** We conducted a two-sample MR analysis using genetic variants associated with gene expression of five Metformin drug targets that also show a genetic association with decreased glycated haemoglobin (HbA1c) levels. Effect sizes are obtained from within UK Biobank for HbA1c, and within AAAgen for AAA risk, a multi-ancestry meta-GWAS analysis of 39,221 cases and 1,086,107 controls.

**Results:** We identified statistically significant evidence of a causal association between a genetic proxy for Metformin action and a decrease in AAA risk, OR=0.58 (95%CI: 0.37-0.90 p=0.015). We estimate that on average a one standard deviation decrease in HbA1c, measured via Metformin gene targets, reduces AAA risk by over 40%.

**Conclusion:** Metformin use in those at increased risk of AAA may reduce incidence of disease. Clinical trials are required to assess the efficacy of Metformin in reducing disease risk.

## Introduction

The only current treatment for abdominal aortic aneurysm (AAA) is surgical repair, which is associated with significant morbidity and mortality. Patients not fit enough for surgery have no treatment options. For people with small AAA there are no treatments to slow AAA growth and prevent the need for surgery in the future.

Diabetes is associated with reduced AAA risk but to date no evidence of a causal association has been found. Observational studies have shown that Metformin, a drug prescribed to lower glycated haemoglobin (HbA1c) in diabetics, is also associated with a reduction in AAA disease risk and reduced growth(1) and may be the driver of the reduced risk seen in people with diabetes. Whilst randomised trials are currently underway to test the effectiveness of metformin to prevent AAA progression(2,3), these are based on observational data and no causal link has been demonstrated. Here we utilise genetic variants to investigate the potential for a causal association between Metformin and AAA risk through Mendelian randomisation (MR).

## Methods

We conducted MR using published instruments of genetic variants for known Metformin targets. Variants were identified through associated expression of genes involved in the action of these targets that also associated with HbA1c levels(4). This resulted in five instruments consisting of a total of 32 variants near 22 genes. The instruments represent Metformin’s HbA1c lowering effect.

Effect sizes were independently estimated in European-ancestry subjects within UK Biobank (n=344,182) and represent changes in Z-standardised HbA1c where a unit change is estimated as 1.09% (∼6.75mmol)(4). This amount is consistent with the effect size estimated in a systematic review of Metformin monotherapy; 1.12% (95%CI: 0.92-1.32)(5).

Effect sizes and standard error estimates for AAA risk were obtained from AAAGen, the largest genetic study on AAA risk to date: a multi-ancestry (European and African) meta-GWAS of 39,214 cases and 1,086,107 controls(6).

MR was conducted using the inverse variance weighted (MR-IVW) method(7) as our primary analysis for each individual target and also using an instrument of the five targets combined. To assess potential weak instrument bias or pleiotropy we applied MR-Egger, weighted-median, weighted-mode, MR-RAPS and MR-PRESSO as sensitivity analyses to the combined instrument(7). Effect estimates were compared across methods for evidence of consistency. To assess the specificity of the Metformin pathway we also assessed a genome-wide HbA1c instrument sourced from the same paper(4).

## Results

Of the 32 independent variants (r^2^<0.01) we found 31 available within AAAgen. Analysing each target revealed consistent protective effects against AAA risk (**Figure** panel **A**). The combined instrument of 31 variants found statistically significant evidence (p=0.015) of a causal association between the HbA1c lowering effects across the five Metformin targets and a decrease in AAA risk. The combined instrument estimates a 43% reduction in risk due to a 1.09% decrease in HbA1c OR=0.57 (95%CI: 0.37-0.90). There was a small amount of overlap between the exposure and outcome samples, as AAAGen included UK Biobank AAA cases and up to 22,049 controls as part of the meta-analysis and the HbA1c effect sizes are estimated within UK Biobank. However, this is anticipated to have minimal effect on the estimates, except for MR-Egger, when using Biobank sized data(8). We also note a potential for increased heterogeneity between the datasets due to mixed ancestry, although allele frequencies between included samples were found to be consistent(6).

**Figure:**
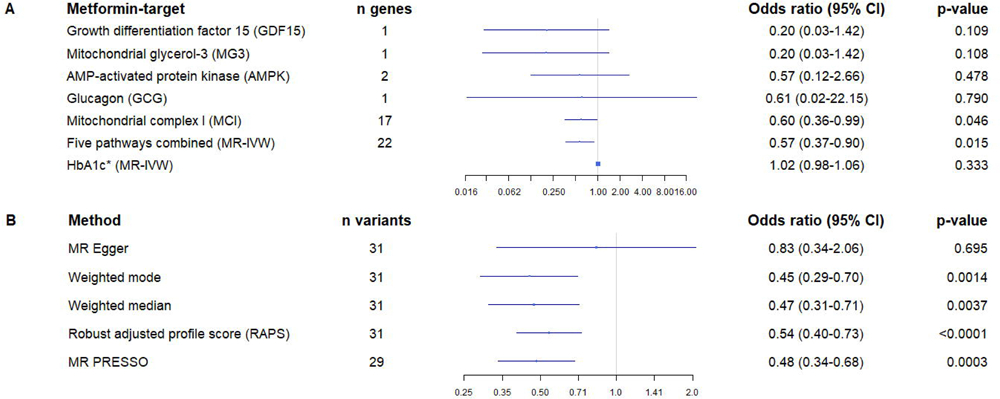
*Non-targeted-whole-genome HbA1c instrument. **Associations between genetically predicted HbA1c levels via 22 genes / 31 variants and abdominal aortic aneurysm risk from two-sample Mendelian randomisation analyses.** Where n genes indicates the number of genes in each pathway. Odds ratio (OR) estimates represent a 1.09% decrease (∼6.75mmol/mol or 1 s.d.) in HbA1c. **Panel A:** Primary analysis of individual Metformin-drug target pathways estimated using MR-IVW, combined pathways of 31 genetic variants in the instrument, and genome-wide HbA1c estimate using inverse weighted variance method showing OR and 95% confidence intervals. **Panel B:** Sensitivity analysis of the combined 31 variant instrument. The MR-PRESSO estimate represents IVW after the removal of two potential outlier variants.

All sensitivity analyses were applied to the combined instrument due to the limited number of genes in each individual target and show strong consistency of effect size and direction (**Figure** panel **B**). The MR-Egger test for a non-zero intercept was not statistically significant (p=0.362) suggesting no evidence of pleiotropy. MR analysis using the genome-wide HbA1c instrument showed no evidence of a causal association with AAA risk, OR=1.02 (95%CI: 0.98-1.06).

## Discussion

This study used genetic instruments to assess the potential therapeutic benefit of Metformin in reducing AAA risk via known Metformin targets. We provide evidence that Metformin has the potential to reduce AAA risk through target-specific HbA1c lowering mechanisms. Results were consistent across targets and sensitivity analyses. The desired reduction in HbA1c to achieve this risk reduction is within the estimated pharmacological effect of a clinical Metformin dose. While we have not shown a direct effect on AAA progression, the inferred benefit of Metformin treatment provides additional justification for ongoing clinical trials (2,3). Critically, no evidence of association with AAA risk was found using a whole-genome HbA1c instrument, giving further evidence to the specificity of an HbA1c change due to Metformin targets.

We acknowledge that the validity of the instrument and its suitability to represent Metformin effects has attracted comment and criticism(9,10). It is highly likely that there are Metformin pathways that have not have been captured here. Other pathways may counteract these protective effects and nullify or, even worse, cause harm and increase AAA risk. Whilst this is possible, it would be inconsistent with findings seen in observational studies. However, we recognise that this paper is limited to reporting results on five known targets and that further work is required to be able to weight these correctly to obtain a more accurate combined effect estimate.

We also note that the instrument was constructed on postulated downstream effects of Metformin on glycaemic levels via known Metformin targets, and we cannot evaluate the effect of Metformin therapy in clinical practice(10). We have so far been unable to identify a suitable and meaningful positive control and therefore cannot confirm that the targets are a representation of the true Metformin effect.

Findings from clinical trials currently in progress to assess the impact of Metformin on AAA growth and risk of serious outcome(2,3) will soon help to shed light on what is difficult to ascertain here: that is, whether these targets represent a valid Metformin instrument or if the effects could be cancelled out by unaccounted pathways. With no medication currently available to reduce AAA risk or slow disease progression, we remain hopeful that our findings, in alignment with those seen in observational studies, will confirm that Metformin may prove to be a viable treatment option.

## Data Availability

All data produced in the present work are contained in the manuscript.

https://eur03.safelinks.protection.outlook.com/?url=https%3A%2F%2Fwww.nature.com%2Farticles%2Fs41588-023-01510-y&data=05%7C01%7Ckls46%40leicester.ac.uk%7C8f991e88d2ac454faf3808dbca7d9f2d%7Caebecd6a31d44b0195ce8274afe853d9%7C0%7C0%7C638326413104819669%7CUnknown%7CTWFpbGZsb3d8eyJWIjoiMC4wLjAwMDAiLCJQIjoiV2luMzIiLCJBTiI6Ik1haWwiLCJXVCI6Mn0%3D%7C3000%7C%7C%7C&sdata=MabkFU6hRMAc0Rd5cDyY6xStDbW2XYK7bne60DgKqDg%3D&reserved=0

## References

1) Golledge J, Moxon J, Pinchbeck J, Anderson G, Rowbotham S, Jenkins J et al. Association between metformin prescription and growth rates of abdominal aortic aneurysms. Br J Surg 2017;104:1486–1493.

2) Lareyre F, Raffort J. Metformin to Limit Abdominal Aortic Aneurysm Expansion: Time for Clinical Trials. Eur J Vasc Endovasc Surg 2021;61:1030.

3) Golledge J, Arnott C, Moxon J, Monaghan H, Norman R, Morris D et al. Protocol for the Metformin Aneurysm Trial (MAT): a placebo-controlled randomised trial testing whether metformin reduces the risk of serious complications of abdominal aortic aneurysm. Current controlled trials in cardiovascular medicine; Trials 2021;22:962.

4) Zheng J, Xu M, Walker V, Yuan J, Korologou-Linden R, Robinson J et al. Evaluating the efficacy and mechanism of metformin targets on reducing Alzheimer’s disease risk in the general population: a Mendelian randomisation study. Diabetologia 2022;65:1664–1675.

5) Hirst JA, Farmer AJ, Ali R, Roberts NW, Stevens RJ. Quantifying the effect of metformin treatment and dose on glycemic control. Diabetes Care 2012;35:446–454.

6) Roychowdhury T. et al. Multi-ancestry GWAS deciphers genetic architecture of abdominal aortic aneurysm and highlights PCSK9 as a therapeutic target. MedRXiv 2022; doi 10.1101/2022.05.27.22275607.

7) Burgess S, Davey Smith G, Davies NM, Dudbridge F, Gill D, Glymour MM et al. Guidelines for performing Mendelian randomization investigations. Wellcome open research; Wellcome Open Res 2019;4:186.

8) Minelli C, Del Greco M F, van der Plaat D A., Bowden J, Sheehan NA, Thompson J. The use of two-sample methods for Mendelian randomization analyses on single large datasets. Int J Epidemiol 2021;50:1651–1659.

9) Anderson EL, Williams DM. Drug target Mendelian randomisation: are we really instrumenting drug use? Diabetologia 2023;66:1156–1158.

10) Burgess S, Mason AM, Grant AJ, Slob EAW, Gkatzionis A, Zuber V et al. Using genetic association data to guide drug discovery and development: Review of methods and applications. Am J Hum Genet 2023;110:195–214.

